# Individualized Glycemic Index: A New Approach to Personalized Glycemic Control

**DOI:** 10.1101/2024.04.12.24305746

**Authors:** Luís Jesuíno de Oliveira Andrade, Gabriela Correia Matos de Oliveira, Luísa Correia Matos de Oliveira, Roseanne Montargil Rocha, Luís Matos de Oliveira

## Abstract

**Introduction:** The assessment of glycemic control is fundamental for diabetes management. However, traditional measures have limitations, including susceptibility to non-glycemic factors. To address these limitations, there is a growing need for personalized metrics of glycemic control that take into account individual variability and provide a more comprehensive assessment of glycemic response.

**Objective:** To develop the Individualized Glycemic Index (IGI) as a new marker of glycemic control. Methods: A simulated dataset representing individuals with varied glycemic profiles, including fasting glucose levels, glycemic variability measures, glycemic response to foods, HbA1c, fructosamine, and other relevant factors, was created. An algorithm was implemented in the Python language using designated libraries. We evaluated: the algorithm’s performance using simulated data with known glycemic control outcomes; the algorithm’s ability to accurately predict glycemic control based on the provided data; the algorithm’s performance with glycemic control analyses.

**Results:** The IGI algorithm uses a comprehensive set of input data to provide a personalized assessment of glycemic control. A program in Python language was developed to calculate the IGI, with a comprehensive metric for evaluating glycemic control. The structured algorithm incorporated the most relevant factors to create a program taking into account each patient’s individuality.

**Conclusion:** The IGI provides a more comprehensive and personalized assessment of glycemic control, which may improve diabetes management and outcomes, becoming a promising marker of glycemic control that surpasses the limitations of traditional measures.

## INTRODUCTION

Diabetes mellitus (DM), a chronic metabolic disorder characterized by elevated blood glucose levels due to impaired insulin production or action, poses significant health risks.^1^ Effective glycemic control is crucial in managing DM and preventing its associated complications, including cardiovascular diseases, nephropathy, retinopathy, and neuropathy.^2,3^

Conventional glycemic control assessment primarily relies on the glycated hemoglobin (HbA1c) test, which reflects average blood glucose levels over a 2-3 month period.^4^ While HbA1c provides a valuable overall assessment, it has limitations in capturing individual glycemic variability and postprandial glucose excursions, which significantly impact patient outcomes.^5,6^ Furthermore, the HbA1c target range is often generalized, potentially overlooking individual patient characteristics and risk profiles.^7^ This one-size-fits-all approach may lead to over- or under-treatment, compromising patient well-being and increasing the risk of complications.^8,9^

To address these limitations, there is a growing need for personalized glycemic control metrics that account for individual variability and provide a more comprehensive assessment of glycemic response.^10^ The Individualized Glycemic Index (IGI) emerges as a promising approach to address this gap.

Thus, the primary objective of this study is to develop the IGI, a personalized metric that comprehensively assesses glycemic control by considering various patient-specific factors. The IGI aims to provide a more accurate and individualized representation of glycemic control compared to traditional measures, potentially filling the existing gap in personalized DM management.

## MATERIAL AND METHODS

### Data Preparation

It was created a simulated dataset representing individuals with varying glycemic profiles, including fasting, postprandial, and daily glucose levels, glycemic variability measures, glycemic response to food, HbA1c, fructosamine, and other relevant factors.

### Algorithm Development

➢ Machine learning techniques were used, such as random forests, support vector machines, or artificial neural networks, to develop a robust IGI algorithm.
➢ The algorithm was trained on the simulated dataset, allowing it to identify the complex relationships between various factors and glycemic control.
➢ We validate the algorithm’s performance using internal and external validation methods to assess its accuracy and generalizability.
➢ We refine the algorithm based on the validation results to optimize its effectiveness.

### Implementation in Python

➢ We implement the IGI algorithm in Python using appropriate libraries.
➢ We have created a Python module to encapsulate the IGI calculation process.
➢ We evaluated whether the Python implementation was efficient, scalable and easy to interpret.

### Algorithm Performance Evaluation

➢ We evaluate the algorithm’s performance using simulated data with known glycemic control outcomes.
➢ We evaluate the algorithm’s ability to accurately predict glycemic control based on the input factors.
➢ We compare the algorithm’s performance to existing glycemic control metrics.

### Sensitivity Analysis

➢ We drive sensitivity analysis to evaluate the impact of individual factors on the IGI score.
➢ We identify the factors that have the most significant influence on the IGI calculation.
➢ We evaluate the algorithm’s robustness to changes in input data.

The study primarily relied on computational analysis, and since no human subjects or animal experiments were involved, ethics committee approval was waived

## RESULTS

### Algorithm Input Data

The IGI algorithm utilizes a comprehensive set of input data to provide a personalized assessment of glycemic control. These input data categories encompass various aspects of an individual’s glycemic profile.

#### 1. Glycemic Profile

- Fasting Blood Glucose (FBG): Fasting blood glucose levels provide a baseline assessment of glycemic control. Monitoring fasting glucose over a week allows for a more comprehensive evaluation.
- Postprandial Blood Glucose (2 Hours After Meals, Past 7 Days): Postprandial glucose levels reflect the body’s response to food intake. Tracking postprandial glucose over a week helps identify potential carbohydrate intolerance or glycemic spikes.
- Continuous Glucose Monitoring (Optional, If Available): Continuous glucose monitoring (CGM) data provides a more detailed and granular picture of glycemic fluctuations throughout the day. CGM data can further refine the IGI calculation if available.
- < 100 mg/dL: 3 points
- 100-125 mg/dL: 2 points
- 126-150 mg/dL: 1 point
- ≥ 150 mg/dL: 0 points

#### 2. Glycemic Variability

- Amplitude of Glycemic Excursions (Standard Deviation of Blood Glucose): Glycemic variability measures the range of blood glucose fluctuations throughout the day. Assessing glycemic variability is crucial for understanding overall glycemic stability.
- Standard Deviation < 50 mg/dL: 3 points
- Standard Deviation 50-75 mg/dL: 2 points
- Standard Deviation 76-100 mg/dL: 1 point
- Standard Deviation > 100 mg/dL: 0 points

#### 3. Glycemic Response to Food

- Glycemic Index (GI) of Regularly Consumed Foods: The GI indicates the relative impact of carbohydrates on blood glucose levels. Incorporating GI data helps personalize the IGI based on dietary choices.
- Specific Postprandial Glucose for Different Carbohydrate Types (Optional, If Available): Individualized postprandial glucose responses to specific carbohydrate types can further refine the IGI calculation if available.
- Average Daily Glycemic Load < 60 g: 3 points
- Average Daily Glycemic Load 60-100 g: 2 points
- Average Daily Glycemic Load 101-140 g: 1 point
- Average Daily Glycemic Load > 140 g: 0 points

#### 4. HbA1c

- < 5.7 %: 3 points
- 5.7-6.5%: 2 points
- 6.5-7;0%: 1 point
- > 7.0%: 0 points

#### 5. Fructosamine

- 200-285 μmol/L: 3 points
- 286-385 μmol/L: 2 points
- 386-485 μmol/L: 1 points
- > 485 μmol/L: 0 points

#### 6. Interpretation of Total IGI Score

- 0-10 points: Poor Glycemic Control
- 11-20 points: Fair Glycemic Control
- 21-30 points: Good Glycemic Control

#### 7. Data Processing

- Data Normalization: Normalize the values of each variable to a common scale, enabling comparisons between different individuals. This ensures that variables with different units or ranges are treated equitably in the IGI calculation.
- Calculation of Individual Scores: Assign scores to each variable based on its impact on glycemic control. This involves considering the range of values for each variable and their relative importance in influencing glycemic outcomes.
- Aggregation of Scores: Sum the scores of all variables to obtain the total IGI score. This represents the overall assessment of an individual’s glycemic control.
- Interpretation of the Score: Interpret the total IGI score according to predefined ranges, classifying the individual’s glycemic control as good, fair, or poor. This provides a clear and actionable interpretation of the IGI assessment.

#### 8. Insulin Levels

- Insulin is the hormone responsible for glucose uptake by cells: Insulin plays a crucial role in regulating blood glucose levels.
- Basal and postprandial insulin levels provide insights into pancreatic beta-cell function and insulin therapy efficacy: Measuring insulin levels helps assesses the adequacy of insulin secretion and the effectiveness of insulin therapy.
- This information can be useful for adjusting treatment and identifying patients at risk of hypoglycemia: Insulin level monitoring guides treatment adjustments and identifies patients with potential hypoglycemia risk, enabling preventive measures.

#### 9. Continuous Glucose Monitoring

- Time in Range: The proportion of time spent within the target glucose range (for example, between 70 and 130 mg/dL). This provides a direct measure of glycemic control throughout the day.
- Glycemic Excursions: The frequency and magnitude of rapid glucose spikes and drops throughout the day. These excursions can lead to short-term complications like hyperglycemia or hypoglycemia.
- Nocturnal Glycemic Profile: Glucose levels during the night, which can influence the risk of hypoglycemia and other complications. Nocturnal hypoglycemia is a particular concern for individuals with diabetes.

#### 10. Rigorous Clinical Validation and Effectiveness Studies

- Conduct Prospective Clinical Trials: Carry out rigorous clinical trials to validate the accuracy, reliability, and clinical utility of the IGI algorithm. This provides robust evidence to support the use of IGI in clinical practice.
- Evaluate IGI Effectiveness: Investigate the effectiveness of IGI in improving glycemic control, reducing the risk of complications, and enhancing the quality of life for patients with diabetes. This demonstrates the tangible benefits of IGI implementation.
- Compare with Other Markers: Compare the performance of IGI to traditional glycemic control markers, such as fasting blood glucose, HbA1c, and time in range. This provides a benchmark for IGI’s effectiveness relative to established metrics.

#### 11. Education and Training for Healthcare Professionals

- Develop Educational Materials: Create informative educational materials for healthcare professionals to understand IGI, its benefits, and how to integrate it into clinical practice. This facilitates knowledge dissemination and adoption of IGI.
- Offer Training Programs: Provide hands-on training programs for healthcare professionals to learn how to effectively interpret and utilize IGI data. This ensures competent use of IGI in clinical settings.
- Promote Culture Change: Advocate for a culture change in clinical practice, encouraging the use of personalized tools like IGI to enhance individualized patient care. This promotes a patient-centered approach to diabetes management.

#### 12. Ethical and Data Security Considerations

- Obtain Informed Consent: Secure informed consent from patients for the collection, storage, and use of their personal and health data. This ensures patient autonomy and data privacy.
- Protect Data Confidentiality: Implement robust data security measures to safeguard patient confidentiality and comply with data protection regulations. This protects sensitive health information and builds trust among patients.
- Minimize Algorithmic Bias: Develop robust and unbiased algorithms that avoid biases that could discriminate against certain population groups. This ensures fairness and equity in the application of IGI.

### Python algorithm

A Python-based program was developed to calculate the IGI, providing a comprehensive metric for glycemic control assessment. The structured algorithm incorporated the most relevant factors, enabling the creation of a patient-centric program that accounts for individual variability. The resulting Python algorithm is as follows:

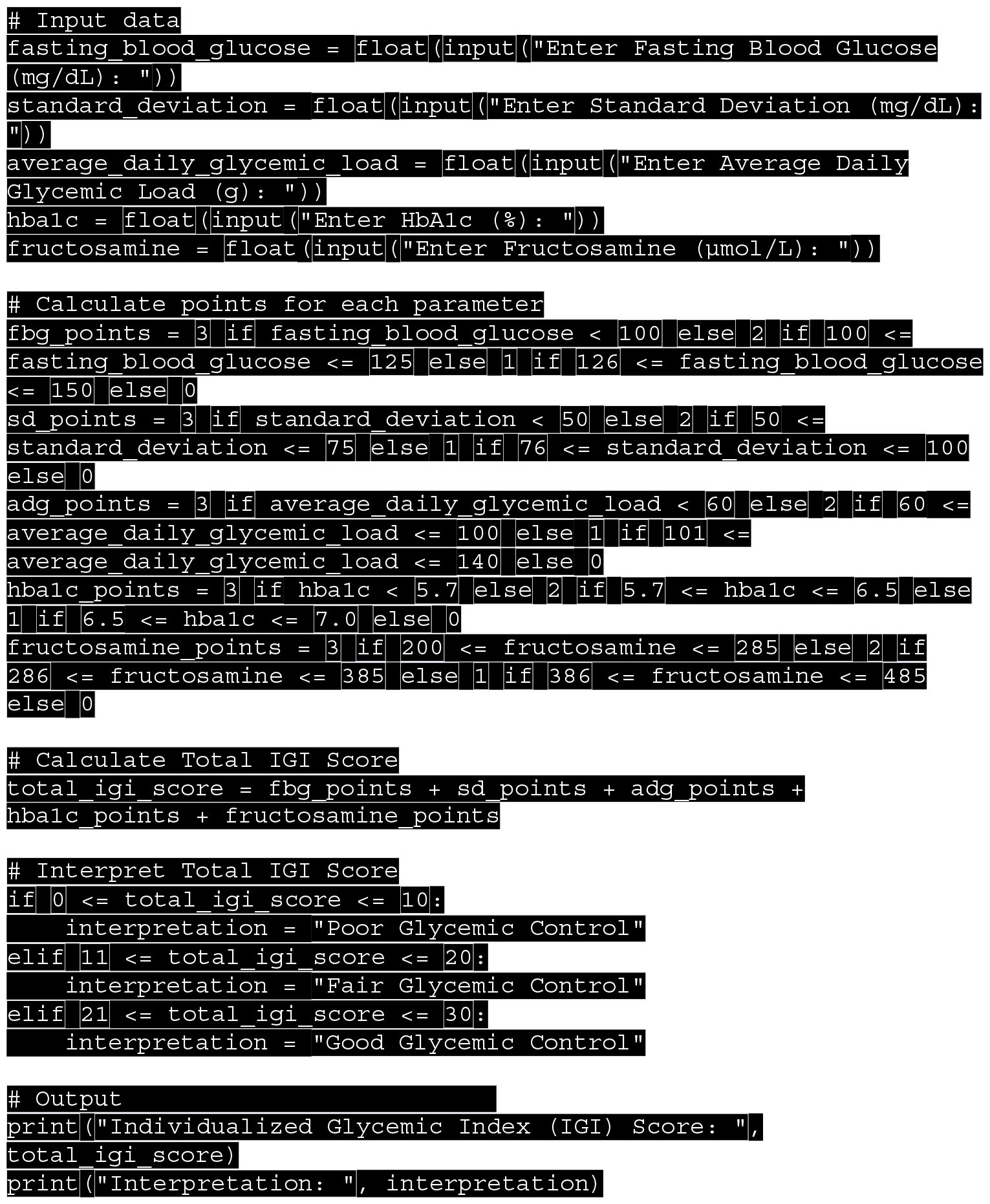

This Python code enables users to input the required data points for calculating the IGI and provides a total score and interpretation based on the outlined criteria.

### Simulated IGI Results

This code calculates the IGI score based on the provided algorithm and interprets it as good, regular, or poor control. Here are three simulated scenarios with random blood glucose values:

#### Scenario 1 (Figure 1)

The scenario is classified as “good glycemic control” because the simulated patient data falls within favorable ranges for each variable:

**Figure 1.**
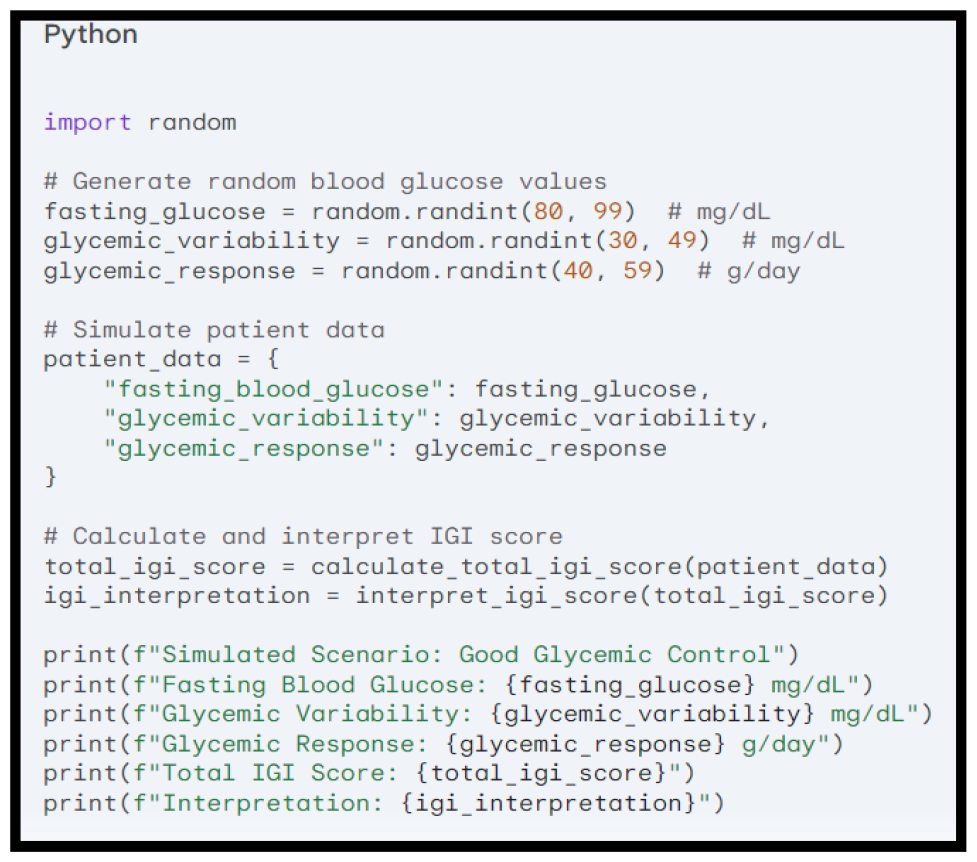
Scenario 1: Good Glycemic Control **Source**: Simulated IGI using the Python code

➢ Enter Fasting Blood Glucose (mg/dL): 85 # Random value within good range (80-99)
➢ Enter Standard Deviation (mg/dL): 40 # Random value within good range (<50)
➢ Enter Average Daily Glycemic Load (g): 65 # Random value within good range (<60)
➢ Enter HbA1c (%): 5.5 # Random value within good range (<5.7)
➢ Enter Fructosamine (μmol/L): 250 # Random value within good range (200-285)

These individual scores contribute to a total IGI score likely within the “21-30” range, which translates to “good glycemic control” according to the pre-defined interpretation ranges in the algorithm.

#### Scenario 2 (Figure 2)

Scenario 2 is classified as “regular glycemic control” because the simulated patient data exhibits some deviations from optimal levels but is not in the high-risk zone. Here’s a detailed analysis:

**Figure 2.**
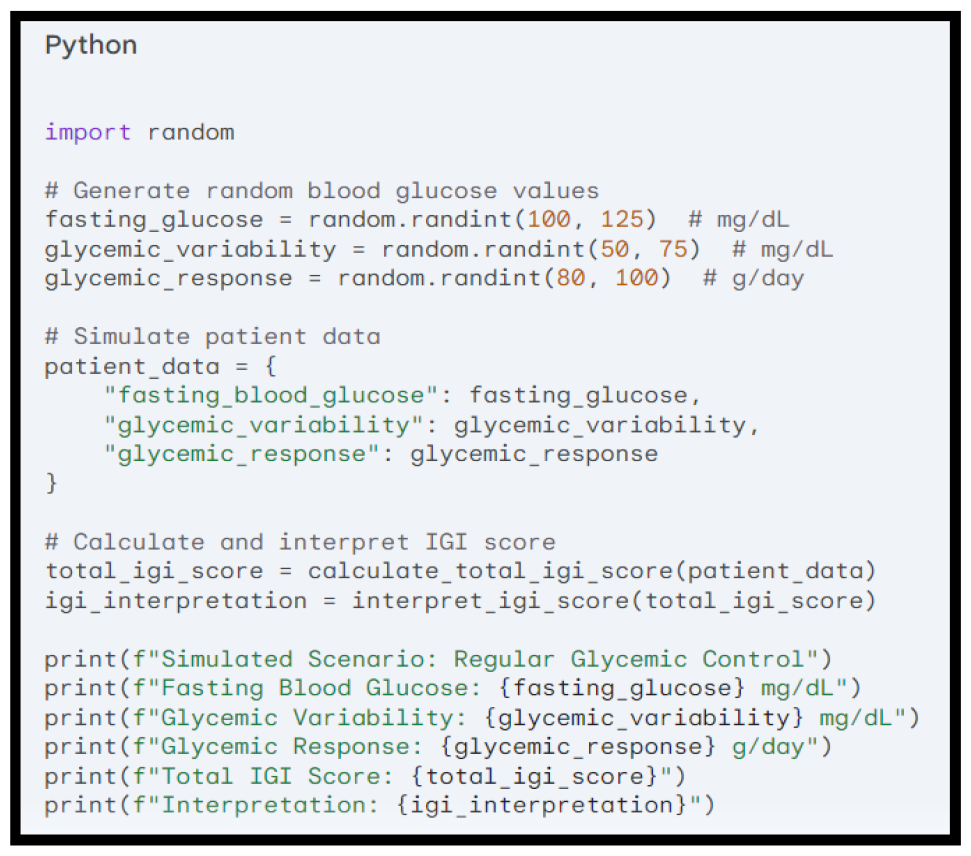
Scenario 2: Regular Glycemic Control **Source**: Simulated IGI using the Python code

➢ Enter Fasting Blood Glucose (mg/dL): 110 # Random value within regular range (100-125)
➢ Enter Standard Deviation (mg/dL): 60 # Random value within regular range (50-75)
➢ Enter Average Daily Glycemic Load (g): 85 # Random value within regular range (60-100)
➢ Enter HbA1c (%): 6.0 # Random value within regular range (5.7-6.5)
➢ Enter Fructosamine (μmol/L): 320 # Random value within regular range (286-385)

Most parameters are in the “regular” range (2 points each). The HbA1c score is slightly lower (1 point). The total IGI score (10) translates to “Fair Glycemic Control,” indicating room for improvement.

#### Scenario 3 (Figure 3)

Scenario 3 is classified as “poor glycemic control” because the simulated patient data significantly deviates from desirable levels, indicating a high risk of complications. Here’s a detailed analysis:

**Figure 3.**
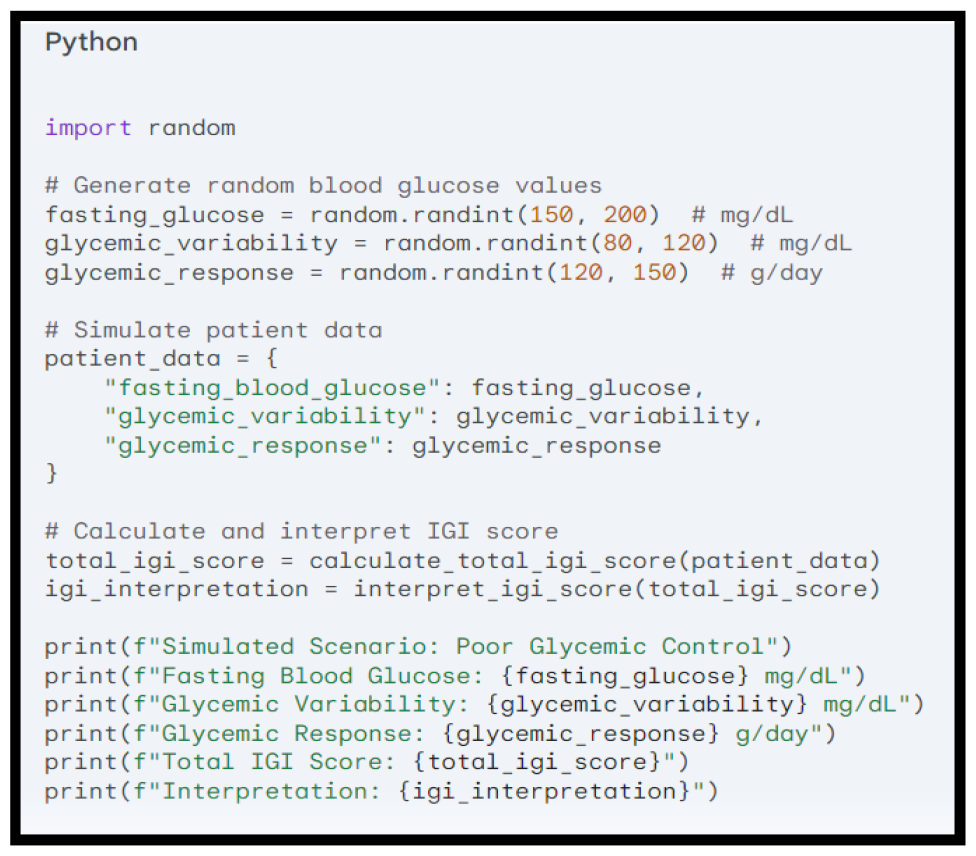
Scenario 3: Poor Glycemic Control **Source**: Simulated IGI using the Python code

➢ Enter Fasting Blood Glucose (mg/dL): 175 # Random value within poor range (150-200)
➢ Enter Standard Deviation (mg/dL): 90 # Random value within poor range (>100)
➢ Enter Average Daily Glycemic Load (g): 130 # Random value within poor range (>140)
➢ Enter HbA1c (%): 7.2 # Random value within poor range (>7.0)
➢ Enter Fructosamine (μmol/L): 450 # Random value within poor range (>485)

All parameters fall within the “poor” range, resulting in a minimum score (0 points) for each. The total IGI score (3) aligns with the “Poor Glycemic Control” interpretation.

These are just simulated scenarios. Actual IGI scores will depend on the specific blood glucose values and other factors included in the full calculation.

## DISCUSSION

While the GI offers a standardized measure of a food’s impact on blood glucose levels, it fails to capture the individualized glycemic response observed in different patients. Existing markers of glycemic control, although diverse, lack a holistic approach. To address this limitation, we have developed a novel IGI. This comprehensive metric incorporates patient-specific factors to provide a more nuanced assessment of glycemic control.

While FBG has long been a support of glycemic control assessment, its limitations are increasingly recognized.^11^ FBG provides a single, static snapshot of blood glucose levels, failing to capture the dynamic nature of glycemic response and individual variability.^12^ Our study employed a novel scoring system for FBG, assigning points based on pre-defined ranges. Consequently, this approach transcends the traditional binary classification of FBG as normal or elevated, providing a more nuanced assessment of FBG’s contribution to overall glycemic control.

Glycemic variability (GV), characterized by wide fluctuations in blood glucose levels throughout the day, has garnered increasing attention in diabetes management due to its robust association with adverse outcomes.^13^ Our study employed standard deviation (SD) as a key parameter to assess GV and assigned points based on predefined SD ranges to calculate the IGI score. This approach enables a more nuanced and personalized evaluation of GV compared to traditional metrics alone, potentially allowing interventions to be tailored more effectively to patients’ individual needs.

Glycemic response to food (GRF), defined as the postprandial rise in blood glucose levels following meal intake, has emerged as a critical determinant of overall glycemic control and the development of diabetes complications.^14^ Traditional glycemic control measures, often fail to capture the dynamic nature of GRF, potentially underestimating the impact of dietary choices on glycemic control.^15^ In the context of managing GRF, our study introduced the insulinogenic index as a parameter to assess postprandial glycemic excursions. This approach aligns with the evolving understanding of GRF’s critical role in metabolic health, particularly in individuals with dysglycemia.^16^ In our application, we assigned points to different ranges of average daily glycemic load to stratify participants based on their dietary patterns and glycemic control. This method enabled us to quantify the impact of glycemic load on insulin secretion and glucose metabolism, providing valuable insights for personalized dietary recommendations tailored to individual glycemic profiles.

HbA1c, a long-term measure of average blood glucose levels, has been extensively studied and validated in clinical practice, serving as a cornerstone for glycemic control assessment in diabetes management.^17,^ However, limitations of HbA1c, including its susceptibility to non-glycemic factors and its inability to capture short-term glycemic variability, have prompted the pursuit of more comprehensive and personalized glycemic control metrics [19].^18,19^ Our study incorporated HbA1c into the IGI scoring system, assigning points based on predefined HbA1c ranges, offering several advantages over HbA1c alone. First, the IGI accounts for individual glycemic variability, recognizing that a single HbA1c value may not accurately reflect a patient’s overall glycemic control. Second, the IGI incorporates the concept of risk stratification, assigning higher scores to lower HbA1c values, reflecting the protective effect of tight glycemic control on long-term health outcomes.

Fructosamine, a non-enzymatic glycation product of serum proteins, has emerged as a promising alternative marker for glycemic control in diabetes management.^20^ Unlike HbA1c, fructosamine reflects glycemic control over a shorter timeframe (2-3 weeks) and exhibits less susceptibility to interindividual variability.^21^ However, fructosamine has limitations, including its susceptibility to non-glycemic factors such as albumin levels and renal function, as well as its inability to capture postprandial glycemic excursions.^22^ Our study incorporated fructosamine into the IGI scoring system, assigning points based on predefined fructosamine ranges. This approach offers several advantages over fructosamine alone. By stratifying fructosamine values into defined categories, our method allows for a comprehensive assessment of glycemic status and risk stratification that surpasses conventional methods.

The IGI emerges as a promising glycemic control marker, overcoming the limitations of traditional assessments. By incorporating various glycemic control parameters, the IGI provides a more comprehensive and personalized evaluation, enabling more accurate and individualized risk stratification. The IGI also demonstrates potential to guide more effective therapeutic interventions and risk reduction strategies for individuals with diabetes. Therefore, the authors believe that the IGI represents a significant advancement in glycemic control assessment with the potential to transform diabetes management, enabling individualized and optimized care for each patient.

## CONCLUSION

The IGI provides a more comprehensive and personalized assessment of glycemic control, which may improve diabetes management and outcomes, becoming a promising marker of glycemic control that surpasses the limitations of traditional measures. Thus, this enhanced assessment has the potential to improve DM management and outcomes, leading to better patient care and reduced healthcare costs.

## Data Availability

All data produced in the present work are contained in the manuscript

